# Time-to-event analysis of factors influencing delay in discharge from a subacute Complex Discharge Unit during the first year of the pandemic (2020) in an Irish tertiary centre hospital

**DOI:** 10.1101/2023.01.03.23284132

**Authors:** Nithya Rajendran, Puteri Maisarah Rameli, Keneilwe Malomo, Declan Byrne, Joseph Browne, Ontefetse Ntlholang

**Author notes:** **Corresponding Author:** Puteri Maisarah Rameli, Department of AMAU/General Internal Medicine, St James’s Hospital, James Street, Dublin 8, Ireland.

## Abstract

**Background:** There is a paucity of data investigating delays in discharges from subacute complex care units exclusively dealing with complex cases. We aimed to analyse factors causing delays in discharges and explore their implications on the healthcare system in terms of length of stay (LOS), during the COVID-19 pandemic, using time-to-event analysis, to assist in future pandemic healthcare planning decisions.

**Methods:** Single-centre, retrospective, cross-sectional study. Demographics, clinical characteristics and length of stay (LOS) of all patients admitted to our subacute Complex Discharge Unit at our model 4 hospital during the pandemic were retrieved from electronic patient records. The outcome variable in our time-to-event analysis was a delay in discharge with a LOS >15 days. A cox proportional hazards regression model delineated factors contributing to delays.

**Results:** 390 patients were admitted between March 2020 - February 2021 to the subacute complex discharge unit. Among these, 326 (83.6%) patients were >65 years of age, and 233 (59.7%) patients were female. The median (IQR) age was 79 (70-86) years, with a median (IQR) LOS of 19.4 (10-41) days. A total of 237 (60.7%) events were uncensored, LOS > 15 days of which 138 (58.2%) were female, 124 (52.32%) had >4 comorbidities, and 153 (39.2%) were censored into LOS ≤15 days and death 19 (4.8%). Kaplan Meier’s plot allowed the comparison of factors causing a delay in discharge to single factors: age, gender, and multimorbidity. A multivariate cox regression analysis adjusted to each strata age, gender and multimorbidity predicted factors affecting LOS: Age strata 65 - 75 & 75 - 85 [HR 0.233; 95% CI (0.077-.708); p=.010] & [HR .301; 95% CI(.155-.588); p<.001] had common factor patient-centred needs prolonging LOS. The common factors complications arising from comorbidities in male [HR .145; 95 % CI (.081 -.261); p<.001] and female [HR .479; 95% CI (.311 -.737); p<.001], and patient-centred needs in male [HR .472; 95 % CI (.243 -.917); p.027] female [HR .361; 95% CI (.215 -.608); p<.001] exhibited statistically significant results.

**Conclusion:** Reducing LOS is imperative for better patient outcomes and hospital management. The most common factors delaying discharge were complexities that arose from primary diagnoses and patient-centred needs such as cognition, psychosocial needs and carer needs. Further research is needed into exploring multimorbidity as a risk factor for mortality in patients who experience prolonged LOS within a complex discharge unit.

## Background

Length of stay (LOS) is the most common outcome measure of effectiveness.[1] It refers to the time expressed in days between admission to and discharges from the hospital.[2] Worldwide, variability in the measure of delayed discharges based on hospital stay exists. Countries like the Netherlands define delayed discharges as LOS exceeding 50% of the average LOS for the general population in the previous calendar year. In countries such as Australia and Singapore, a delayed discharge is defined as LOS exceeding 21 days and 35 days, respectively. A notional application of the Dutch Model to the hospital inpatient enquiry (HIPE) dataset provides estimates of possible delayed discharge but does not define delayed LOS days in Ireland.[3] During the COVID-19 pandemic, the average inpatient LOS was 5.8 days, representing a slight increase in the average inpatient LOS of 5.7 days between 2016–2019.[2]

Tackling delays in patient discharge is an ongoing challenge, and with the emergence of the COVID-19 pandemic, there has been a greater strain on the Irish healthcare system. Subacute Complex Discharge Units (CDU) are a subset of intermediate care that provides person-centred planning[4] through an individualised, integrated care pathway to improve functional outcomes and reduce LOS.[5] Other factors to consider are defined under Delayed transfers of Care (DTOC): delayed discharges awaiting discharge home, community care provision through the Home Support Service, or placement or transfer to long-term residential places. Formerly, a delayed discharge was described as a patient deemed medically fit for discharge from an acute bed who remained in the hospital because they were awaiting support or care following discharge.[6] However, the COVID-19 pandemic has caused disruptions in discharge planning, discontinuity, and incoordination between subacute CDU and community care,[4] due to service curtailment and limited step-down care options.[7]

There is a paucity of data investigating discharge delays from a subacute CDU dealing with patients with ongoing complex care needs.[8] Multimorbidity (MM) is a term used to define the presence of 2 or more chronic medical conditions in an individual<u>[9]</u>. Harrison et al further elaborate complex MM as the co-occurrence of 3 or more chronic medical diagnoses involving 3 or more systems in the body.[10] Frailty[11] and MM are common causes of delayed discharge among older adults leading to poor functional outcomes. There is also a significant correlation between cognitive impairment and dependency on overall LOS.[12] Moreover, patient-centred needs comprising of cognitive dysfunction, psychosocial needs, carer involvement, and community services linking Home Support Services have also been described.[5] Prolonged hospital stays could harm patient health, for example, through deconditioning, increased risk of healthcare-associated infections (HCAI), and mortality.[2]

Furthermore, delayed discharges have implications on healthcare costs. According to the Health Service Executive (HSE) Performance report 2020, DTOC was notable in 11.6% of patients with complex needs. Furthermore, 5.2% of patients had rehabilitation needs leading to delays in the transfer of care. In 2020, there were 32 reported cases of hospital-acquired Staphylococcus aureus bloodstream infection and 58 cases of Clostridium difficile infection.[2]

This study aims to analyse factors causing delays in discharge from a subacute CDU during the first year of the COVID-19 pandemic.

## Methods

### Setting

The study was conducted in St James’s Hospital (SJH), the largest acute academic teaching model 4 hospital in Ireland, based in the south inner city of Dublin. Within SJH, there is a 23 bedded CDU, which specialises in coordinating the safe and effective discharge of medically stable patients. Through early goal-oriented input from multi-disciplinary team members, it is envisaged that the LOS within post-acute CDU does not exceed six weeks. This retrospective, cross-sectional study provides valuable insight into factors contributing to delayed discharges from a subacute CDU, especially during the first year of the COVID-19 pandemic.

### Data sources

All information was sourced directly from the hospital database via electronic patient records (EPR). In particular, we focused on all adult patients admitted to the CDU, between March 2020 - February 2021 of the pandemic, with hospital LOS>15 days. Two independent reviewers extracted and evaluated the suitability of all data. Information collected was stratified by age, gender, primary diagnoses, the prevalence of MM, corresponding LOS, and causes for delayed discharges. MM was categorised using the Charlson comorbidity index - where 0 was assigned to no comorbidity and 1 for each comorbidity. All details pertaining to delayed discharges were extracted directly from inpatient notes and medical discharge summaries.

### Statistical analyses

All statistical analyses were conducted using IBM SPSS, version 29. Comparative analysis of LOS between March 2020 - February 2021 was performed using Kaplan Meier for each factor: age, gender, MM, and common reasons for DTOC. Further analyses with COX regressions using multiple covariates were used to identify factors causing delays during the period examined. Patients were compared and analysed by survival analysis, the Kaplan-Meier method was used for single-factor comparison, and the Cox Regression Model was used for multi-factor comparison. The event of interest coded with the value of 1 in our study was delayed discharge; those with LOS<15 days and death were assigned a value of 0 (right censored events). Our main objective was to predict a delay in discharge. With the standard cox regression model, we encountered violations of proportional hazard. Therefore, we used a stratified Cox model allowing for covariates with non-proportional hazard, stratum: age, gender, and MM.

## Results

A total of 390 patients were admitted between March 2020 - February 2021 to the CDU. Among these, 326 (83.6%) patients were >65 years of age with a median age (IQR) of 79 (70-86) years. The gender distribution of men and women was 40.3% vs 59.7%, respectively, 188 (48.20%) had > 4 comorbidities. For our study, we defined prolonged LOS as a hospital LOS >15 days. During the COVID-19 pandemic, the CDU median (IQR) LOS was 19.4 (10-41) days. The most common primary diagnosis on admission was infection or sepsis (23.33%). Each stratum was preliminarily analysed using the Mann-Whitney test and chi-square (Gender) respectively. (Table 1)

**Table 1:** Strata qualitatively assessed using Mann Whitney and Chi-square (Gender) respectively for patients admitted to the complex discharge unit.

|  | 2020 (n=390) |  |  |
| --- | --- | --- | --- |
| Patient Characteristics | LOS ≤15 days<br>(n=153) | LOS >15 days<br>(n = 237) | P <0.05<br>Statistical<br>significance |
| Age (Mean ± SD) | 75.25 ± 12.50 | 77.51 ± 12.62 | p=.038 |
| Gender:<br>Female<br>Male | 62.09%<br>37.90% | 58.22%<br>41.77% | p= 0.51<br>(yates correction to<br>avoid type 1 error);<br>p 0.46 |
| Multimorbidity:<br>≤4<br>>4 | 15<br>64 | 99<br>124 | p <0.05 |

Following this, we proceeded with time-to-event analysis. Kaplan Meier’s plot allowed the comparison of factors causing a delay in discharge to single factors: age, gender, and MM. A total of 237 (60.7%) events were uncensored, LOS > 15 days of which 138 (58.2%) were female, 124 (52.32%) had >4 comorbidities, and 153 (39.2%) were censored into LOS ≤15 days and death 19 (4.8%).

The Kaplan-Meier method allowed a comparison of five major factors responsible for delays in discharge from the CDU: complications arising from primary diagnoses or MM, HCAI, frailty or falls necessitating integrated rehabilitation, patient-centred needs, and community services against LOS.

In our study, more female than male patients experienced delays in LOS. (Fig. 1B) The mean age of patients with delayed LOS was 77.51 years compared to those with normal LOS with a mean age of 75.25 years. Patients between 65-75 years of age were more likely to experience prolonged LOS. (Fig. 1A) Furthermore, patients with >4 MM were most likely to experience a delay in LOS exceeding >15 days. (Fig. 1C) Complications arising directly from comorbidities were a non-contributory factor in prolonging LOS >15 days. The most common complications arising from comorbidities were cardiovascular: arrhythmias (15.8%) and Left Ventricular dysfunction (20%), hypertension (25.3%), neurological: cognitive impairment secondary to infection (17.2%), worsening of Parkinson’s disease (8.78%) and respiratory: exacerbation of airway disease (4.8%). Endocrine complications accounted for 8.1% of patients with diabetes mellitus and thyroid disorders. (Fig. 1D) Patients whose inpatient course was complicated by HCAI were not observed to have prolonged LOS >15 days. (Fig. 1E) Additionally; frailty, falls and/or integrated rehabilitation or community services did not result in prolonged LOS >15 days. (Fig.1F, H) Patient-centred needs were a significant contributing factor to prolonged LOS >15 days. (Fig. 1G) Our univariate Km plot analysis showed statistically significant results p<.05 for age and factors such as complications that arose from comorbidities, frailty or falls necessitating integrated rehabilitation and patient-centred needs.

**Fig. 1.**
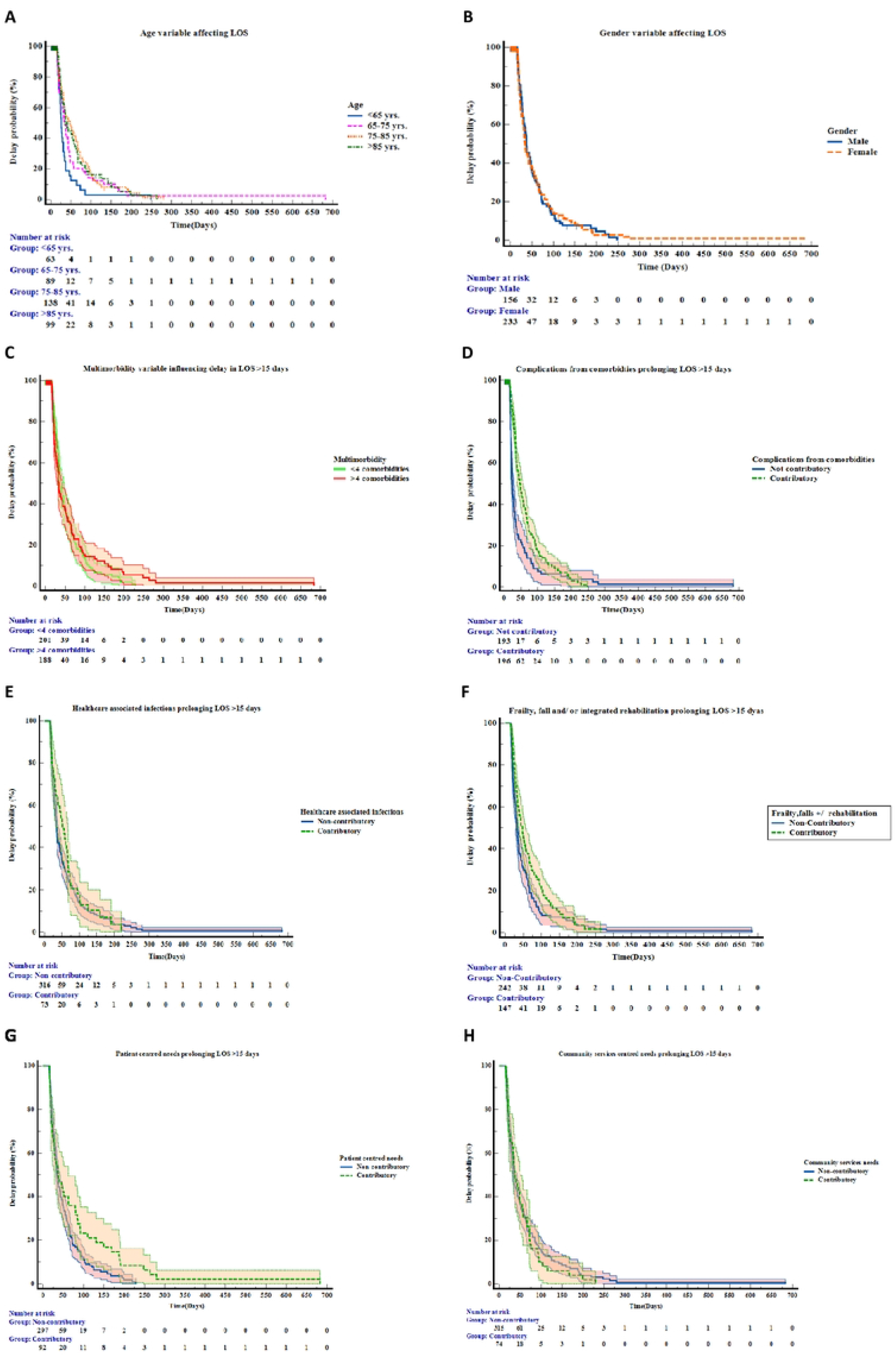
Kaplan-Meier delay in discharge plots of each variable. **A)** Patients between 65 -75 years of age were more likely to experience prolonged LOS. **B)** Patients of the female gender were observed to have prolonged LOS in comparison to their male counterparts. **C)** Patients with >4 MM were most likely to experience a delay in LOS exceeding >15 days. **D)** Complications arising directly from comorbidities are a non-contributory factor in prolonging LOS >15 days. The most common complications arising from comorbidities were cardiovascular: arrhythmias (15.8%) and Left Ventricular dysfunction (20%), hypertension (25.3%), neurological: cognitive impairment secondary to infection (17.2%), worsening of Parkinson’s disease (8.78%) and respiratory: exacerbation of airway disease (4.8%). Endocrine complications accounted for 8.1% of patients with diabetes mellitus and thyroid disorders. **E)** Patients whose inpatient course was complicated by HCAI were not observed to have prolonged LOS >15 days in comparison to those who did not acquire HCAI. **F)** Frailty falls and/or integrated rehabilitation did not result in prolonged LOS > 15 days. **G)** Patient-centred needs were a significant contributing factor to prolonged LOS >15 days. **H)** Community services were not a significant factor leading to prolonged LOS > 15 days.

Using stratified COX regression analysis, we analysed age, gender and MM against the five factors associated with delay in discharge. (Table 2) The five main factors discussed were not responsible for delayed LOS in patients >85 years. In a comparison of each stratum to the five main factors causing a delay in discharge, it was noted that complications arising from primary diagnoses and patient-centred needs had statistically significant results p <.05.

**Table 2:** Variables independently associated with delay in discharge (LOS >15) from Stratified COX regression analysis.

| Chief Characteristics |  | Covariates | B Coefficient | Hazard Ratio (95% CI) | P Value |
| --- | --- | --- | --- | --- | --- |
| Age | <65 | Complications/comorbidities prolonging discharge | -2.329 | .097<br>(.028 -.340) | <.001 |
|  |  | Healthcare-associated infection | -1.005 | .366<br>(.095-1.416) | .146 |
|  |  | Frailty, falls and/or integrated rehabilitation needs | -.063 | .939<br>(.272-3.239) | .921 |
|  |  | Patient-centred needs | -.039 | .962<br>(.325-2.850) | .944 |
|  |  | Community Services | -.216 | .806<br>(.321-2.020) | .645 |
| Age | 65 - 75 | Complications/comorbidities prolonging discharge | -.759 | .468<br>(.206-1.065) | .070 |
|  |  | Healthcare-associated infection | .190 | 1.209<br>(.506-2.887) | .669 |
|  |  | Frailty, falls and/or integrated rehabilitation needs | -.893 | .410<br>(.211-.795) | .008 |
|  |  | Patient-centred needs | -1.455 | .233<br>(.077-.708) | .010 |
|  |  | Community Services | 0.67 | 1.069<br>(.522-2.188) | .855 |
| Age | 75 -85 | Complications/comorbidities prolonging discharge | -1.270 | .281<br>(.160-.492) | <.001 |
|  |  | Healthcare-associated infection | -.628 | .533<br>(.288-.987) | .045 |
|  |  | Frailty, falls and/or integrated rehabilitation needs | -.473 | .623<br>(.387-1.002) | .051 |
|  |  | Patient-centred needs | -1.199 | .301<br>(.155-.588) | <.001 |
|  |  | Community Services | -.075 | .928<br>(.536–1.605) | .789 |
| Age | > 85 | Complications/comorbidities prolonging discharge | -.619 | .539<br>(.272-1.065) | .075 |
|  |  | Healthcare-associated infection | -.177 | .838<br>(.369-1.904) | .673 |
|  |  | Frailty, falls and/or integrated rehabilitation needs | -.213 | .808<br>(.427-1.531) | .514 |
|  |  | Patient-centred needs | -.498 | .608<br>(.262-1.408) | .246 |
|  |  | Community Services | -.203 | .817<br>(.394-1.693) | .586 |
| Gender | Male | Complications/comorbidities prolonging discharge | -1.930 | .145<br>(.081-.261) | <.001 |
|  |  | Healthcare-associated infection | -.574 | .563<br>(.314-1.010) | .054 |
|  |  | Frailty, falls and/or integrated rehabilitation needs | -.235 | .790<br>(.513-1.217) | .286 |
|  |  | Patient-centred needs | -.751 | .472<br>(.243-.917) | .027 |
|  |  | Community Services | -.027 | .973<br>(.621-1.524) | .905 |
| Gender | Female | Complications/comorbidities prolonging discharge | -.736 | .479<br>(.311-.737) | <.001 |
|  |  | Healthcare-associated infection | -.157 | .854<br>(.524-1.393) | .528 |
|  |  | Frailty, falls and/or integrated rehabilitation needs | -.521 | .594<br>(.401-.880) | .009 |
|  |  | Patient-centred needs | -1.018 | .361<br>(.215-.608) | <.001 |
|  |  | Community Services | -.073 | .930<br>(.520-1.663) | .809 |
| Multi morbidity | ≤4 | Complications/comorbidities prolonging discharge | -.858 | .424<br>(.267-.672) | <.001 |
|  |  | Healthcare-associated infection | -.351 | .704<br>(.411-1.207) | .202 |
|  |  | Frailty, falls and/or integrated rehabilitation needs | -.843 | .431<br>(.285-.651) | <.001 |
|  |  | Patient-centred needs | -.828 | .437<br>(.247-.772) | .004 |
|  |  | Community Services | -.234 | .792<br>(.501-1.252) | .317 |
| Multi morbidity | >4 | Complications/comorbidities prolonging discharge | -1.179 | .308<br>(.190-.497) | <.001 |
|  |  | Healthcare-associated infection | -.086 | .917<br>(.551-1.528) | .740 |
|  |  | Frailty, falls and/or integrated rehabilitation needs | -.256 | .774<br>(.512-1.170) | .224 |
|  |  | Patient-centred needs | -1.045 | .352<br>(.192-.644) | <.001 |
|  |  | Community Services | .196 | 1.211<br>(.739-2.002) | .442 |

This analysis predicted factors affecting LOS: Age strata 65 - 75 & 75 - 85 [HR 0.233; 95% CI (0.077-.708); p=.010] & [HR .301; 95% CI (.155-.588); p<.001] had common factor patient-centred needs prolonging LOS. Interestingly, both genders and individuals with ≤4 MM and >4 MM had complications arising from primary diagnoses and patient-centred needs prolonging LOS with p <.05, statistically significant results. The common factors complications arising from comorbidities in male [HR .145; 95 % CI(.081 -.261); p<.001] and female [HR .479; 95% CI (.311 -.737); p<.001]; and patient-centred needs in male [HR .472; 95 % CI (.243 -.917); p.027] female [HR .361; 95% CI (.215 -.608); p<.001] exhibited statistically significant results.

## Discussion

This retrospective cross-sectional analysis of patients admitted within the first year of the COVID-19 pandemic to a subacute CDU delineated five factors responsible for delays in discharge. First, it was evident that the COVID-19 pandemic resulted in many patients exceeding average LOS in hospital with subsequent impact on transfers of care: 37.94% of patients experienced LOS exceeding >15 days as a result of complications arising from primary diagnoses or MM, 25.38% due to frailty or falls necessitating integrated rehabilitation, 14.35% consequent to issues surrounding community services provision, 13.07% due to patient-centred needs and 10.52% due to HCAI.

When comparing our patient demographics to the current literature, we noted that the distribution of age and gender did not vary significantly pre and during the COVID-19 pandemic. Many patients admitted to our CDU were female, predominantly over 65 years of age, and affected by three factors common to delaying LOS within their age group. These factors were complications arising from primary diagnoses or MM, frailty necessitating integrated rehabilitation, and patient-centred needs.[13] Our findings were similar to a recent Singaporean study that showed how continuity of integrated care in the community was considerably affected during the COVID-19 pandemic. In their study, reluctance or hesitancy from formal and informal caregivers to accept community services due to fear of contracting COVID-19 infection resulted in many patients experiencing delayed LOS.[8] Another study from the US made similar observations when evaluating discharge processes in a skilled nursing facility.[14] The pressured situation observed in social care, especially long-term care facilities, is something that all hospitals are internationally familiar with and continuously struggle to find resolutions.

We acknowledge that the occurrence of MM is associated with higher rates of admission to the hospital and subsequent readmission.[15] Through the utilisation of the Charlson comorbidity index scoring, we sub-categorised complex MM as follows: ≤4 as mild to moderately complex and those with 5 or more as highly complex MM. [16] In our analysis, we were able to ascertain the association of MM with LOS. Complications arising directly from comorbidities were a non-contributory factor in prolonging LOS >15 days. Interestingly, patients admitted to the CDU with ≤4 MM and >4 MM experienced the same delaying factors for LOS, including complications arising from primary diagnoses or MM and patient-centred needs. Further research is needed into exploring MM as a risk factor for mortality in patients who experience prolonged LOS within a CDU.

### Strengths and Limitations

To our knowledge, this is the first study that has investigated the time-to-event analysis of factors influencing delayed discharges, exclusively in complex patients admitted to a CDU during the COVID-19 pandemic. The EPR system used for the study allowed for ease in traceability and transparency of all data. Nevertheless, there were some limitations to the study. Our study was conducted in a single centre, explicitly focusing on delaying factors as opposed to factors leading to readmission. The study solely focused on patients admitted to the CDU instead of incorporating all hospital admissions and readmissions during the COVID-19 pandemic. Third, the interpretation of our findings is limited to the sample size; therefore, the extrapolation and application of data may differ across other centres. Finally, mortality and readmission were not explored in detail during this study.

## Conclusion

Reduced length of stay is imperative for better patient and hospital outcomes. The most common factors delaying discharge were complexities that arose from primary diagnoses and patient-centred needs such as cognition, psychosocial needs and carer needs. The analysis of these factors associated with delays in discharge during the pandemic can aid in medical decisions and form the framework for future contingency planning on resources should we encounter another pandemic.

## Data Availability

All relevant data are within the manuscript and its Supporting Information files.

## Abbreviations

CDU: Complex discharge unit
DTOC: Delayed transfers of care
EPR: Electronic Patient Record
LOS: length of stay
MM: multimorbidity

## Declarations

### Ethical approval and consent to participate

Approval was sought and granted by both the SJH Research & Innovation Office and St James’s/Tallaght University Hospital Research & Ethics Office before conducting the research. Informed consent and a data protection impact assessment (DPIA) were not required as no identifiable information was collected for the study.

### Consent for publication

Not applicable

### Availability of data and materials

All data relevant to the study are included in the article or uploaded as supplementary information

### Author’s contributions

NR and PMR - Conceptualisation, data collection, interpretation, design analysis, drafting, revising and approval of the final manuscript. ON and JB - Data collection. DG, JB, KM and ON - Contributed to the data collection, revising and approval of the final manuscript,

## Acknowledgements

Not applicable

## Supplementary Information

**Additional File 1: Figure S1.**
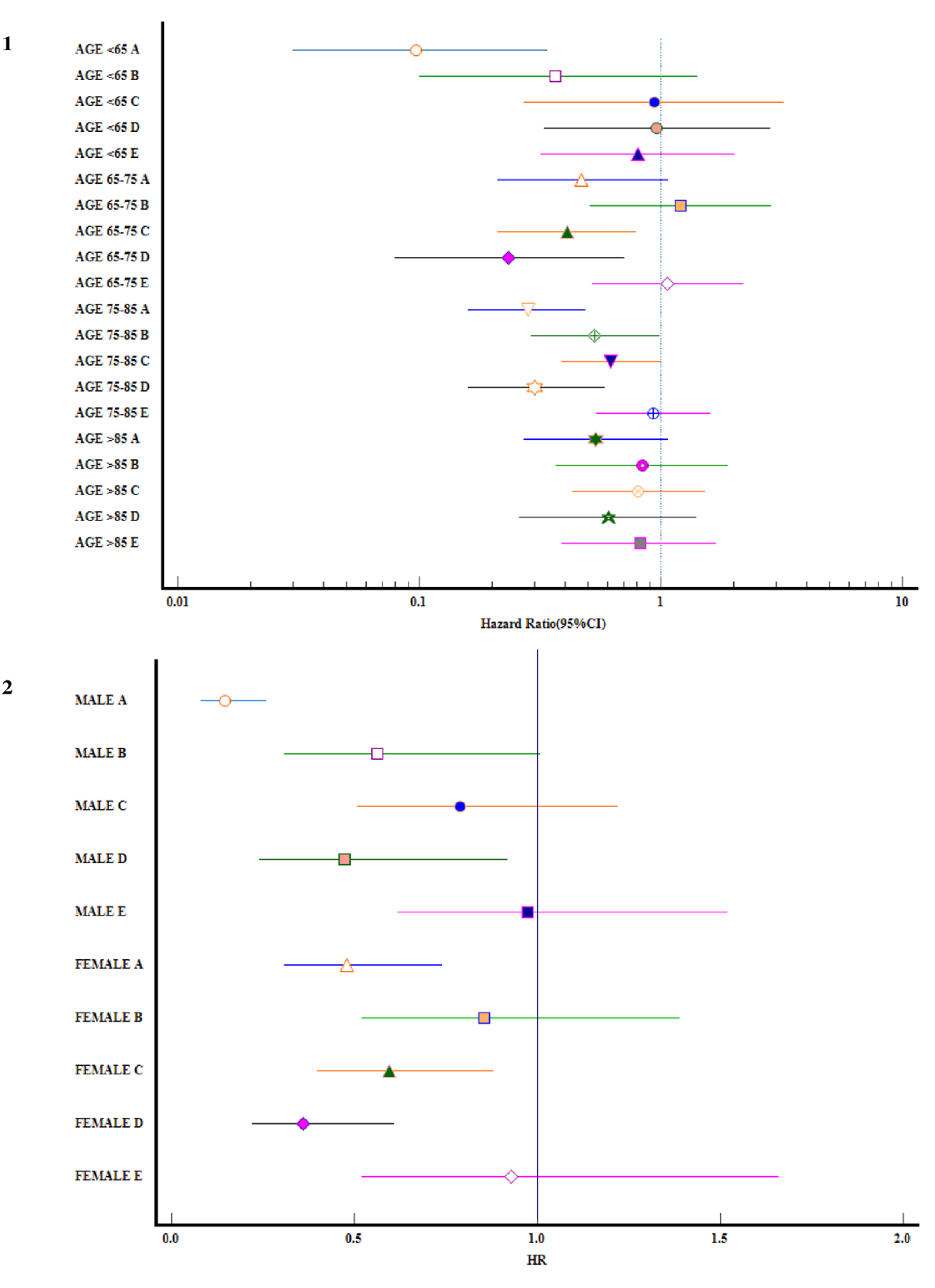
Forest plots 1 and 2 depicting age and gender strata associated Hazard ratio (Markers) estimates (95% Confidence Interval demonstrated by horizontal line) exhibited statistically significant results for individuals < 65 years of age who had a delay in discharge due to complications from comorbidities; those in 65-75 years of age category, had prolonged LOS due to admission with frailty, falls and/or integrated rehabilitation needs; and 75-85 years of age category showed an association of at least 4 out of the 5 common delaying factors. Strata Gender exhibited a significant delay in discharge due to complications from comorbidities and patient-centred needs; in comparison to the female gender who also experienced a delay in discharge as a result of both factors alongside frailty, falls and/or integrated rehabilitation needs.**[A**. Complications/comorbidities prolonging discharge, **B**. Healthcare-associated infection, **C**. Frailty, falls and/or integrated rehabilitation needs, **D**. Patient-centred needs, **E**. Community services]

**Additional File 2: Figure S2.**
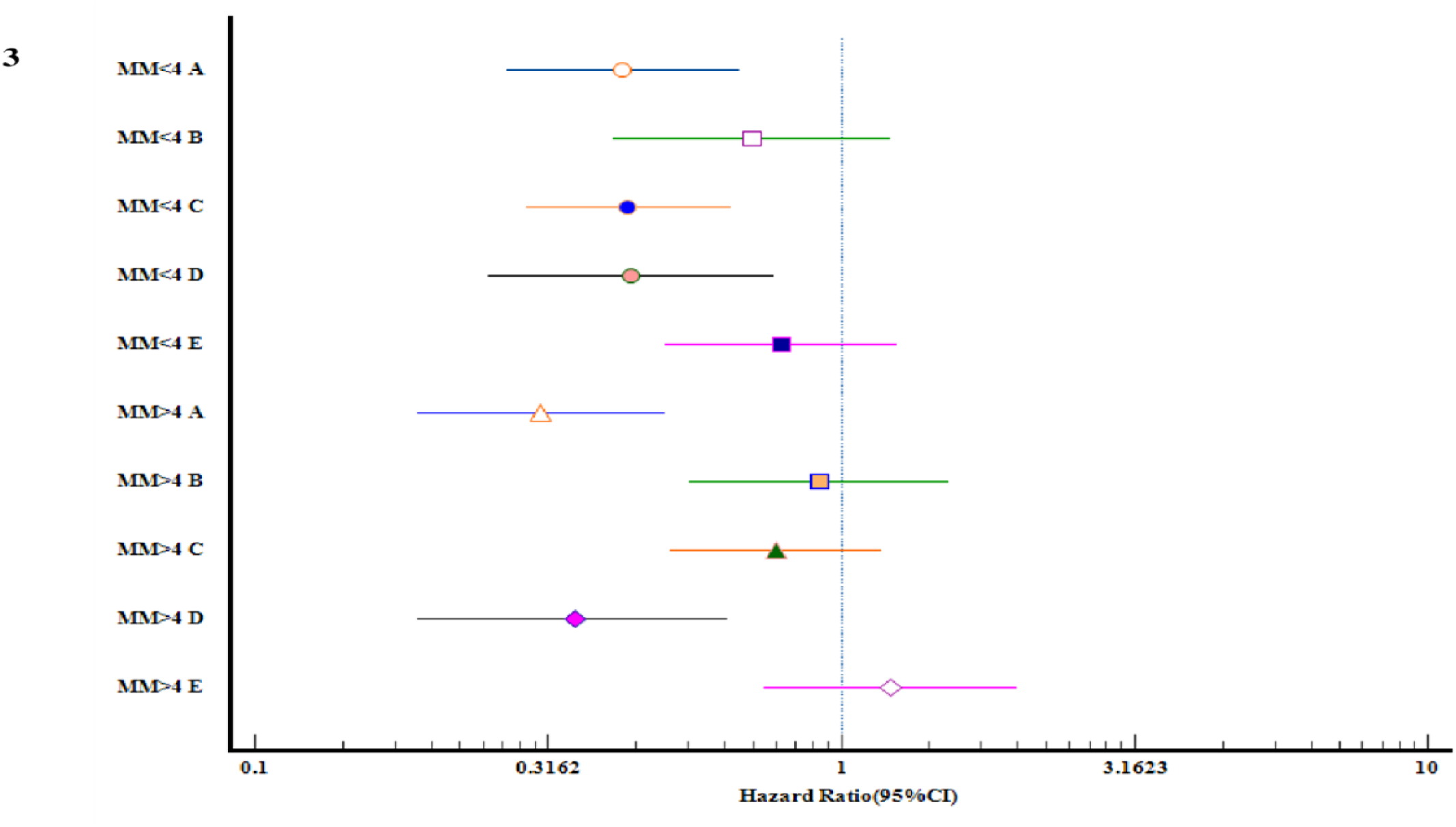
Forest plot 3 depicting multimorbidity (MM) strata-associated Hazard ratio (Markers) estimates (95% Confidence Interval demonstrated by horizontal line) exhibited a significant delay in discharge due to complications from comorbidities, frailty, falls, and/or integrated rehabilitation and patient-centred needs in patients with ≤4 MM. In contrast patients with >4 MM experienced significant delays in discharge due to complications from comorbidities and patient-centred needs.**[A**. Complications/comorbidities prolonging discharge, **B**. Healthcare-associated infection, **C**. Frailty, falls and/or integrated rehabilitation needs, **D**. Patient-centred needs, **E**. Community Services]

